# Effect of Iron-Containing Micronutrient Powders on Growth and Malaria-Induced Anaemia Among Preschool Children in Ghana: A Cluster-Randomized, Double-Blind, Placebo-Controlled Trial

**DOI:** 10.64898/2026.04.24.26351649

**Authors:** Emmanuel Kwesi Yeboah Tchum, Joana Edem Koto, Fred Kanyoke, Ophelia Opoku, Francis Ussher, Slyvester Donnie Dassah, Benjamin Amoani, Samuel Kofi Tchum, Eric Nyarko

## Abstract

**Background:** Affecting 40% of infants and young children worldwide, anaemia in sub-Saharan Africa hampers cognitive and physical development, often in ways that cannot be reversed. Iron-based micronutrient powders (MNPs) are recommended to combat anaemia, but concerns remain about their safety and effectiveness in malaria-endemic areas. We evaluated the impact of iron-based MNPs on growth measurements and malaria-related anaemia among preschool children in Ghana.

**Methods:** We conducted a secondary analysis of a cluster-randomized, double-blind, placebo-controlled trial in the Bono Region, Ghana. Children aged 6–35 months (n=1,958) received daily MNP containing 12·5mg elemental iron or placebo for five months. Anthropometric indices, haemoglobin, and malaria parasitaemia were assessed at baseline and endline. Adjusted analysis of covariance (ANCOVA) models estimated effects on height-for-age (HAZ), weight-for-age (WAZ), and weight-for-height (WHZ) z scores. Binomial regression with identity link estimated risk differences for malaria-induced anaemia. Cluster-robust standard errors were applied at the compound level, and intracluster correlation coefficients (ICCs) were estimated.

**Results:** 1,815 (92·7%) children completed the endline survey, but 1,806 were included in the final analysis. Baseline characteristics were balanced between groups. Iron-containing MNP had no significant effect on endline HAZ (β=0·026, p=0·609), WAZ (β=–0·015, p=0·719), or WHZ (β=–0·035, p=0·463). However, the intervention reduced the risk of malaria-induced anaemia (risk difference 0·050, 95% CI 0·004–0·096; p=0·032). Female sex was associated with higher HAZ (β=0·149, p=0·005).

**Conclusion:** Iron-containing MNP did not improve short-term growth but was associated with a modest reduction in malaria-induced anaemia. These findings support the safe use of iron fortification in malaria-endemic settings while underscoring the need for integrated strategies to address persistent growth faltering and gender specificity.

**TRIAL REGISTRATION clinicaltrials.gov Identifier:** NCT01001871. Registered 27/10/2009, http://www.ClinicalTrials.gov/NCT01001871.

## Introduction

Anaemia and undernutrition remain major contributors to child morbidity and mortality globally, disproportionately affecting infants and young children in sub-Saharan Africa (1, 2). The causes are multifactorial, driven by the dual burden of malaria and nutritional deficiencies, especially iron deficiency (ID) (3, 4). Iron deficiency (ID) and malaria often overlap as major causes of anaemia and poor growth, posing a difficult biological and public health dilemma, made more delicate because giving iron supplementation can inadvertently fuel malaria parasites by increasing the amount of circulating iron available to them (5, 6). Universal iron supplementation strategies, including multiple micronutrient powders (MNPs), are recommended by the World Health Organization (WHO) in high-prevalence areas (7, 8). Although MNPs are recommended by the WHO to reduce anaemia, their use in malaria-endemic settings has been controversial since the Pemba trial reported increased adverse outcomes with iron supplementation (9, 10). This controversy has led to cautious global guidelines and hesitancy regarding programmes (11). Subsequent evidence suggests that the safety and effectiveness of iron interventions depend on formulation, dose, and the context of malaria control (7).

Beyond its high prevalence, paediatric anaemia exerts a profound toll by severely impairing cognitive development and physical growth, often with lasting or irreversible consequences (12, 13). However, whether iron fortification improves growth outcomes while maintaining safety in high-malaria-transmission settings remains uncertain. Additionally, the effects of iron-containing MNPs on gender are not well understood, and it is also unclear whether they may make children more vulnerable to growth faltering. This distinction is important because anthropometric improvements and reductions in anaemia may follow different biological pathways (14). Therefore, this study aimed to assess the effect of iron-containing multiple micronutrient powders on child growth and malaria-induced anaemia among preschool children, while evaluating regression residual distributions across models and groups to ensure model adequacy and identify potential violations of key assumptions in a high-transmission setting in Ghana

## Methods

### Study Design and Participants

This was a secondary analysis of data obtained on the 17^th^ June 2025 from a community-based, double-blind, placebo-controlled, cluster-randomized trial conducted from March to September 2010 in Wenchi Municipal and Tain District, Bono Region, Ghana, an area with perennial, high-intensity *Plasmodium falciparum* transmission (15). The original trial assessed the effect of MNPs on malaria morbidity (16). The present analysis focuses on anthropometric and haematological outcomes.

Children aged 6–35 months were eligible if they were consuming weaning foods, afebrile, free from major illness, and residing in the study area (16, 17). Exclusion criteria included severe anaemia (haemoglobin [Hb] <7 g/dL), severe wasting (weight-for-height Z-score [WHZ] <-3), oedema, congenital abnormalities, recent iron supplementation, or any chronic illness. Ethical approval for the original trial was obtained from the relevant institutional review boards and was subsequently amended as necessary, in accordance with approved procedures, as described in detail elsewhere (18, 19). Written informed consent was obtained from parents or guardians (Caretakers) in the presence of a potential witness as previously described elsewhere (17).

### Randomization and Blinding

Compounds with at least one eligible child formed a cluster. Clusters were randomly assigned (1:1) using a computer-generated sequence to receive either MNP sachets with iron (Intervention) or without iron (Placebo). Sachets were identical in appearance and taste, marked only with a code (‘A’ or ‘B’). Participants, caregivers, field staff, and researchers were blinded to group allocation. The randomization code was held by the manufacturer and research pharmacists and was revealed only to the Data and Safety Monitoring Committee (DSMC) if significant safety concerns arose, and to researchers after database lock, as described elsewhere (15). The authors did not have access to any information that could identify individual participants at any stage of the retrospective study, either during data collection or thereafter, as all data were anonymized.

### Intervention

The intervention group received a daily MNP sachet containing 12.5 mg of elemental iron (as microencapsulated ferrous fumarate), 5 mg of zinc, 400 μg of vitamin A (as retinol equivalent), 30 mg of vitamin C, and other vitamins and minerals. The placebo sachet was identical except that it did not contain iron. Both were to be mixed with a small amount of food once daily for five months. Field researchers conducted weekly home visits to monitor adherence, collect morbidity data, and distribute sachets as mentioned in detail elsewhere (15).

### Procedures

At baseline and endline (5 months), capillary blood samples were collected for Hb measurement (HemoCue, Angelholm, Sweden), full blood count (Horiba ABX Micros 60-OT, France), and preparation of blood smears for malaria microscopy. Malaria parasitaemia was determined by light microscopy of Giemsa-stained thick and thin films as described in detail elsewhere (17). Anaemia was defined as Hb <11 g/dL (15). Malaria-induced anaemia was defined as the presence of both malaria parasitaemia and anaemia. Anthropometric measurements (weight, length/height) were taken by trained personnel using calibrated equipment (SECA scales and stadiometers, Hamburg, Germany) following WHO protocols (15, 19). Z-scores for HAZ, WAZ, and WHZ were calculated using the WHO 2006 Growth Standards (18, 19). Clustering was accounted for using cluster-robust standard errors specified at the compound level. Intracluster correlation coefficients (ICCs) were estimated for each outcome, and design effects were calculated to quantify the impact of clustering on variance estimates.

### Statistical Analysis

All analyses were conducted in R (version 4.3.0, New Zealand) and followed a prespecified analysis plan. Baseline characteristics were summarised using appropriate descriptive statistics. The primary analysis followed the intention-to-treat principle, including all participants with available endline data. For continuous outcomes, the dependent variables were endline anthropometric indices: height-for-age Z score (HAZ), weight-for-age Z score (WAZ), and weight-for-height Z score (WHZ). These were analysed using Analysis of Covariance (ANCOVA) models with the independent variable of interest specified as treatment allocation (intervention vs control). Models were adjusted for prespecified covariates, including the corresponding baseline Z score and child sex, and robust standard errors were used to account for potential heteroscedasticity.

For binary outcomes, the outcome variable was malaria-induced anaemia at endline. This was analyzed using a generalized linear model (GLM) with a binomial distribution and identity link function to estimate adjusted risk differences, with treatment allocation as the primary independent variable and child sex included as a prespecified covariate. Model assumptions and adequacy were assessed using prespecified regression diagnostics. For continuous outcomes analyzed using linear regression, linearity was evaluated using residual-versus-fitted plots, normality of residuals was assessed using quantile–quantile plots, and homoscedasticity was examined using scale–location plots. These diagnostics indicated no substantial deviations from model assumptions, supporting the appropriateness of the fitted models.

All statistical tests were two-sided, and a p-value of less than 0.05 was considered statistically significant. Clustering at the compound level was accounted for using robust standard errors with clustering specified at the cluster level. Where appropriate, intracluster correlation coefficients were estimated to assess the degree of within-cluster similarity.

## Results

### Study Population

A total of 2,220 children from 1,716 clusters across 22 communities in Wenchi Municipal and Tain District were screened for eligibility. 262 children (from 241 clusters) were excluded before randomization. The main reasons for exclusion were failure to meet the inclusion criteria (n=200) and absence during the recruitment period (n=62) (Figure 1). Overall, 1,958 children aged 6–35 months from 1,549 clusters were enrolled and randomly assigned to receive either an iron MNP or a placebo. 967 children were allocated to the iron MNP group and 991 to the placebo group (Figure 1). 1,815 children (92.7%) completed the endline survey, comprising 900 children (93.1%) from 740 clusters in the iron MNP group and 915 children (92.3%) from 723 clusters in the placebo group. After further exclusions, 1,806 children were included in the final analysis, comprising 894 participants in the iron MNP group and 912 in the placebo group. Among the 67 children lost to follow-up in the intervention group, the most common reasons were temporary absence (n=43), relocation from the study area (n=17), and unknown reasons (n=4); three deaths were recorded. Similarly, in the placebo group, among the 76 who did not complete the endline survey, the main reasons were temporary absence (n=46), relocation (n=22), and unknown reasons (n=6); two deaths occurred during follow-up (Figure 1). However, the cause of death among the five participants was not related to the trial, as described in detail elsewhere (20).

**Figure 1:**
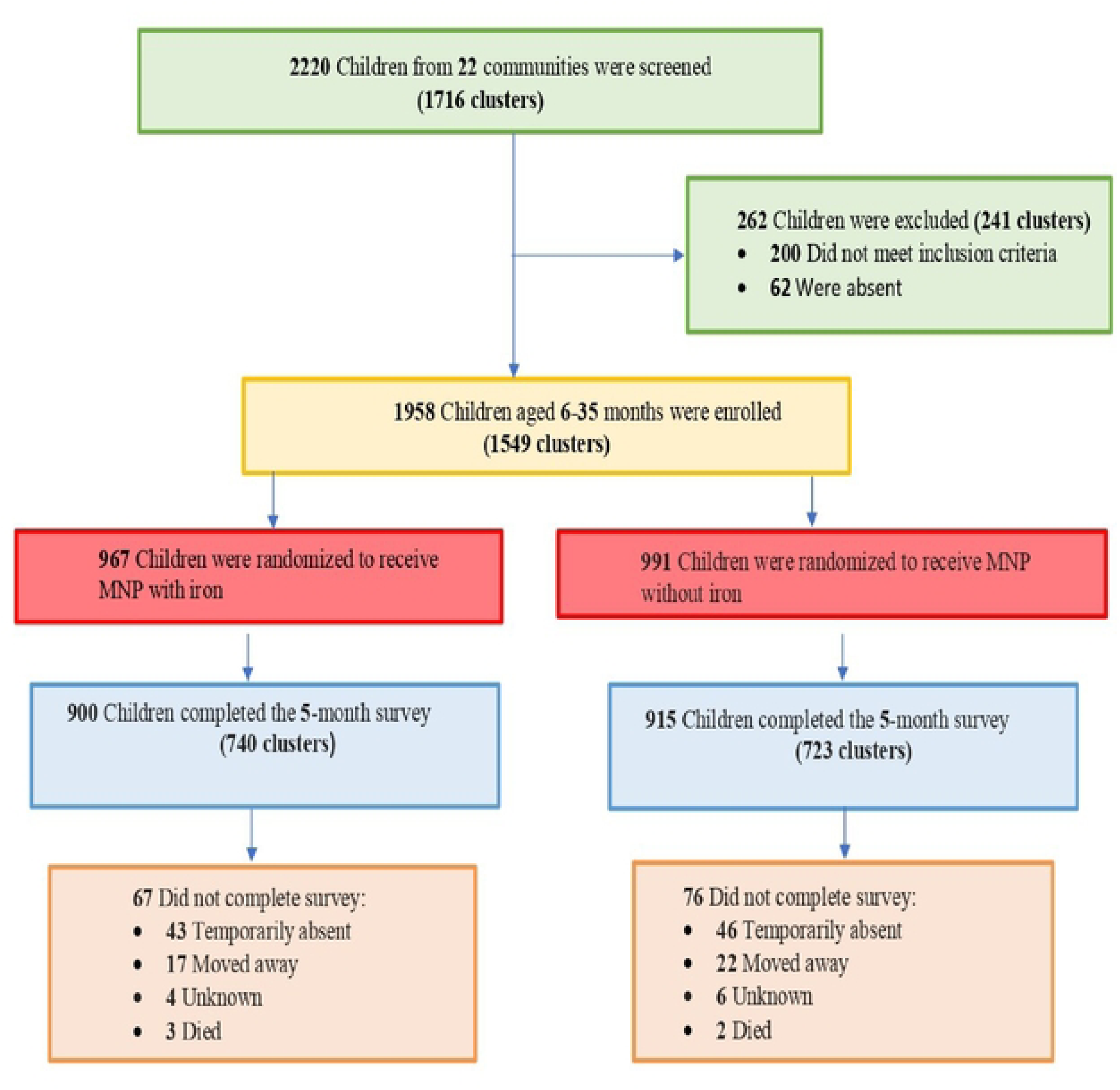
Study profile.

### Background Characteristics among Study Participants

Baseline characteristics of the study participants were generally similar between the intervention and placebo groups (p > 0.05) (Table 1). The mean age of enrolled children was 19.3 months (SD 8.7) in the iron MNP group, and 19.1 months (SD 8.6) in the placebo group (p=0.660). The distribution of sex, household characteristics, and baseline nutritional indicators showed no significant differences between the groups (p>0.05).

**Table 1:**
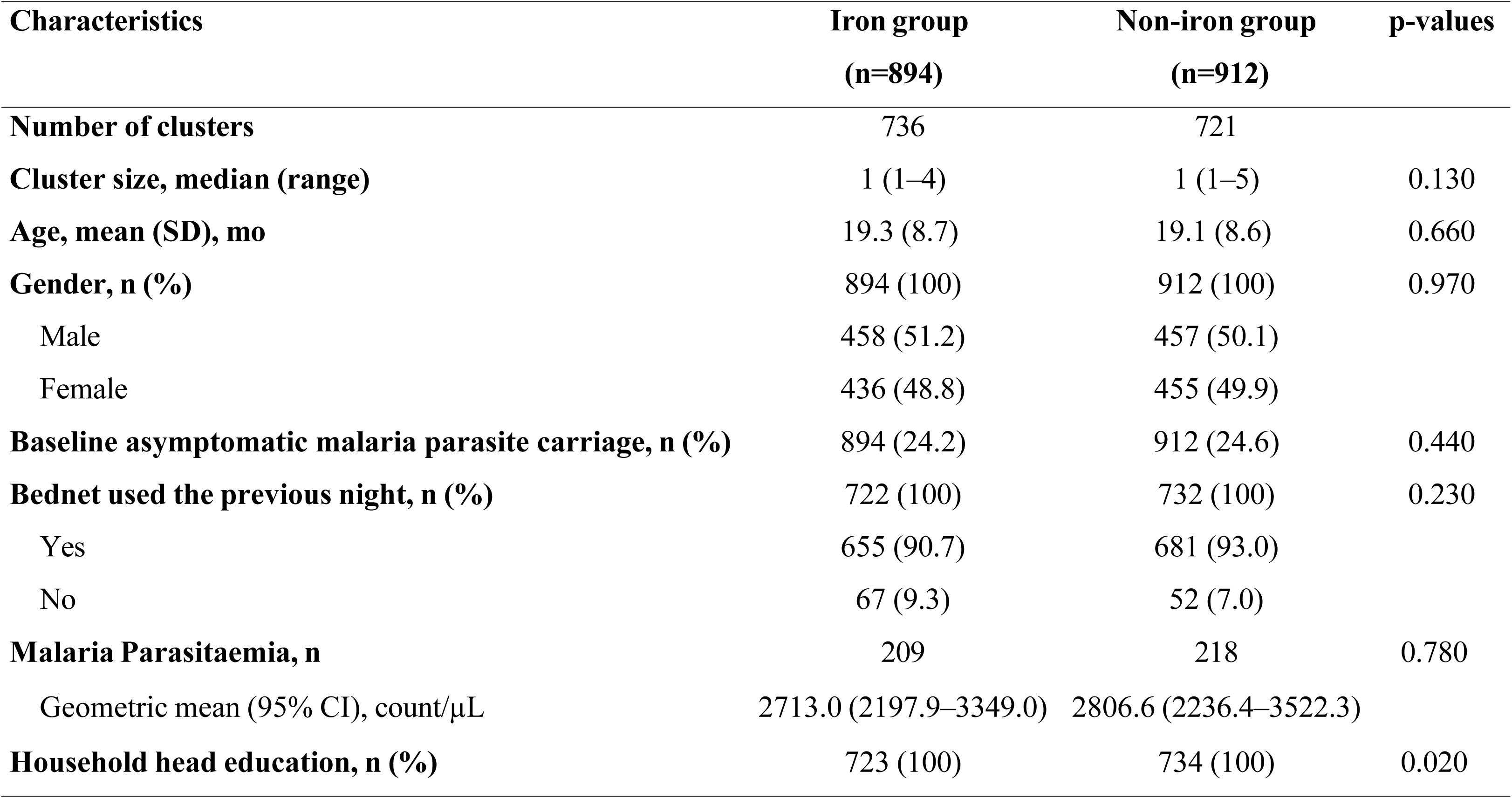
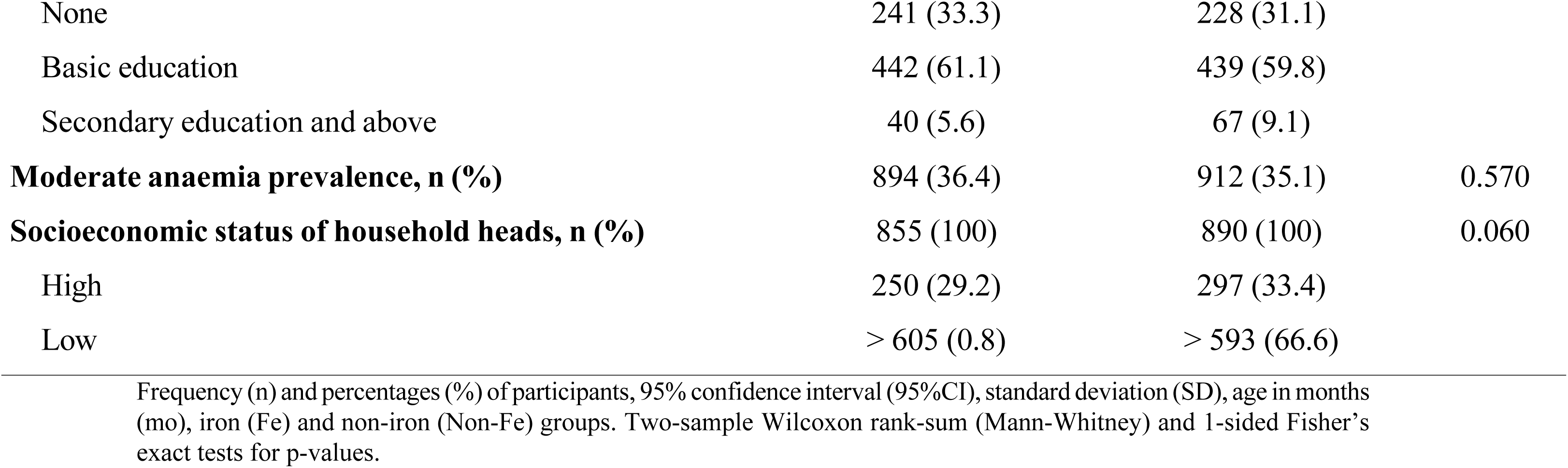
Background characteristics among study participants.

At enrolment, the prevalence of stunting, wasting, and underweight was similar across both groups (p > 0.05). In the iron MNP group, stunting was observed in 14.8% of children, wasting in 8.4%, and underweight in 12.9%. In the placebo group, stunting was present in 13.7%, wasting in 8.0%, and underweight in 12.1%. Baseline mean anthropometric z scores were also comparable between groups (p > 0.05) (Table 1).

The prevalence of baseline malaria parasitaemia and anaemia did not differ significantly between the intervention and placebo groups (p > 0.05), indicating successful randomisation. Other sociodemographic and economic variables, including caregiver education, household size, and community distribution, were well balanced across groups (p > 0.05). The baseline prevalence of asymptomatic malaria (∼24%), moderate anaemia (∼36%), and malnutrition indicators was similar across groups (p > 0.05) (Table 1).

### Nutritional Status of Study Participants

At baseline, stunting prevalence was 14.8% in the iron MNP group and 13.7% in the placebo group (p=0.520). Wasting prevalence was 8.4% and 8.0%, respectively (p=0.760), while underweight prevalence was 12.9% and 12.1% (p=0.620). Chi-square tests confirmed no statistically significant between-group differences at baseline for stunting (χ²(3)=0.78, p=0.855), wasting (χ²(3)=6.55, p=0.088), or underweight (χ²(3)=1.27, p=0.736). At endline, stunting prevalence increased substantially in both groups to 29.2% in the iron MNP group and 27.7% in the placebo group (p=0.490). Wasting declined to 2.5% and 2.3% (p=0.870), and underweight declined to 8.7% and 8.1% (p=0.680). Between-group chi-square tests at endline likewise showed no significant differences for stunting (χ²(3)=2.80, p=0.424), wasting (χ²(3)=3.50, p=0.321), or underweight (χ²(3)=1.62, p=0.654) (Figure 2).

**Figure 2:**
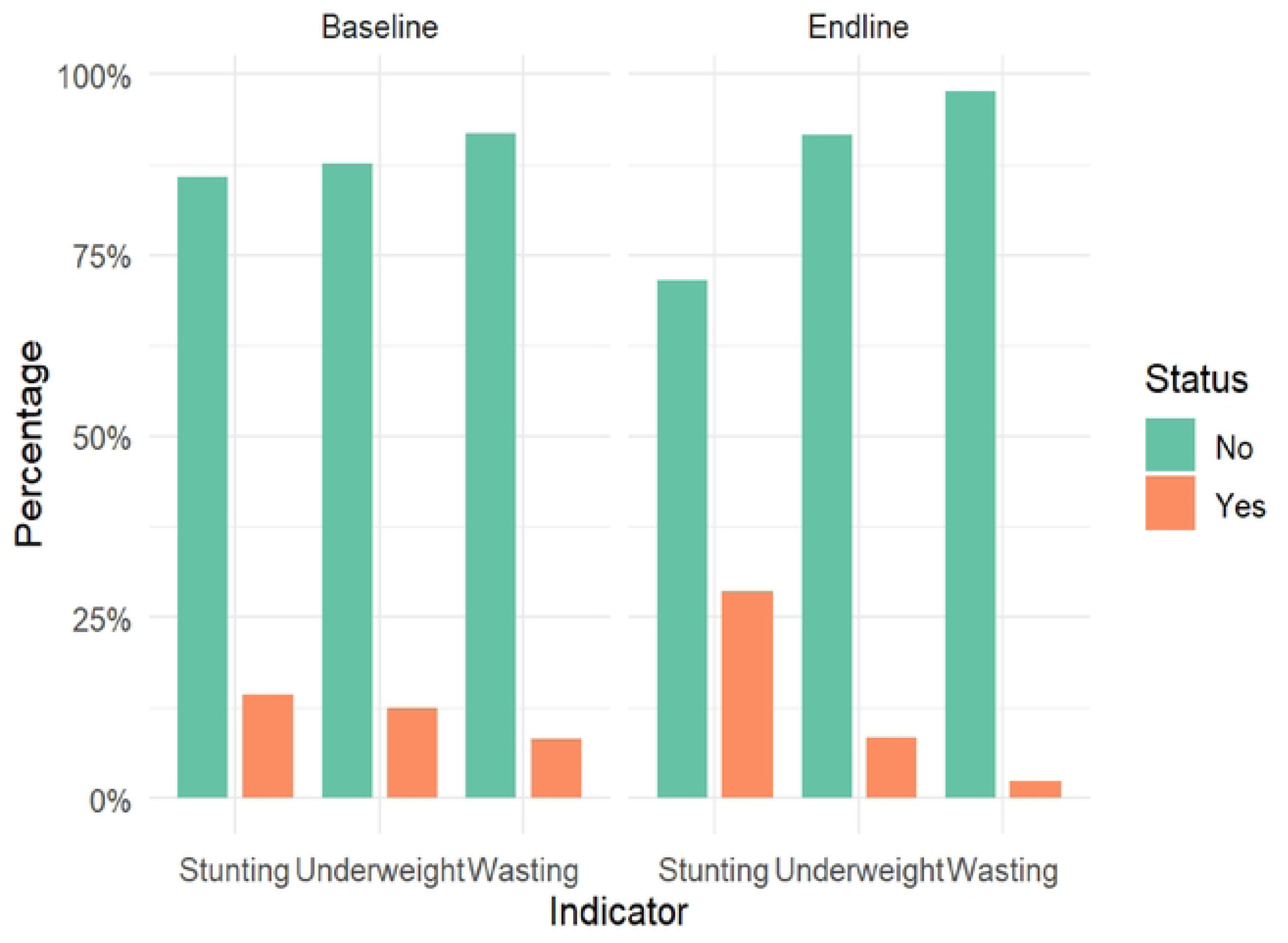
Malnutrition Indicators: Baseline vs Endline.

Within both groups, Wilcoxon signed-rank tests showed statistically significant changes in all three nutritional indicators from baseline to endline (p<0.001 for all comparisons). Wasting and underweight improved significantly in both the iron MNP and placebo groups, whereas stunting worsened markedly in both arms. The parallel nature of these changes, with no significant between-group differences, suggests that secular trends or shared environmental influences, rather than the iron MNP intervention itself, drove the observed shifts in acute malnutrition. The concerning rise in stunting prevalence in both groups highlights the persistence of chronic growth faltering irrespective of supplementation, suggesting differential effects on chronic versus acute malnutrition (Figure 2).

### Effect of iron-containing micronutrient powder on anthropometric outcomes

Across all three anthropometric indicators, iron-containing MNP supplementation was not significantly associated with improvements in growth outcomes compared with placebo (Table 2). For example, the WAZ showed an estimated treatment effect of β = –0.015 (95% CI –0.097 to 0.067; p = 0.719). This coefficient indicates that children receiving iron-containing MNP had a marginally lower mean WAZ at endline than the non-iron group, although the difference was very small and statistically non-significant. The confidence interval includes zero, meaning the true effect could range from a slight decrease to a small increase in WAZ (Table 2). Similarly, the HAZ showed an insignificant association with linear growth. The estimated effect size was β = 0.026 (95% CI: –0.074 to 0.126; p = 0.609), indicating a very small and statistically non-significant rise in HAZ among children receiving iron-containing MNP (Table 2). Again, the confidence interval spans zero, showing no clear evidence of an intervention effect on stunting or linear growth during the study period (Table 2). However, the WHZ, which reflects acute nutritional status or wasting, was insignificantly different between the groups (Table 2). The estimated coefficient was β = –0.035 (95% CI: –0.129 to 0.059; p = 0.463), indicating a small, insignificant decrease in WHZ among children receiving iron-containing MNP (Table 2). In summary, these results suggest that daily iron-containing MNP fortification during the study period did not produce measurable improvements in anthropometric indicators among preschool children in this malaria-endemic setting.

**Table 2:**
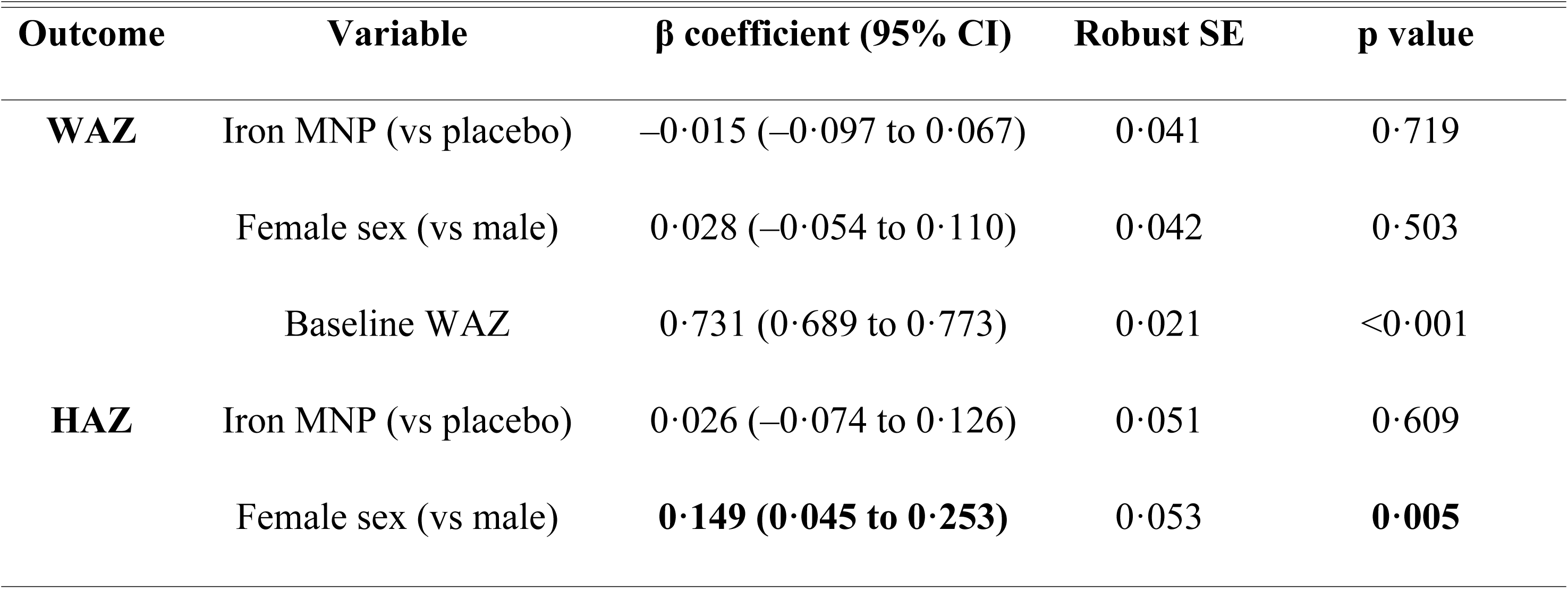
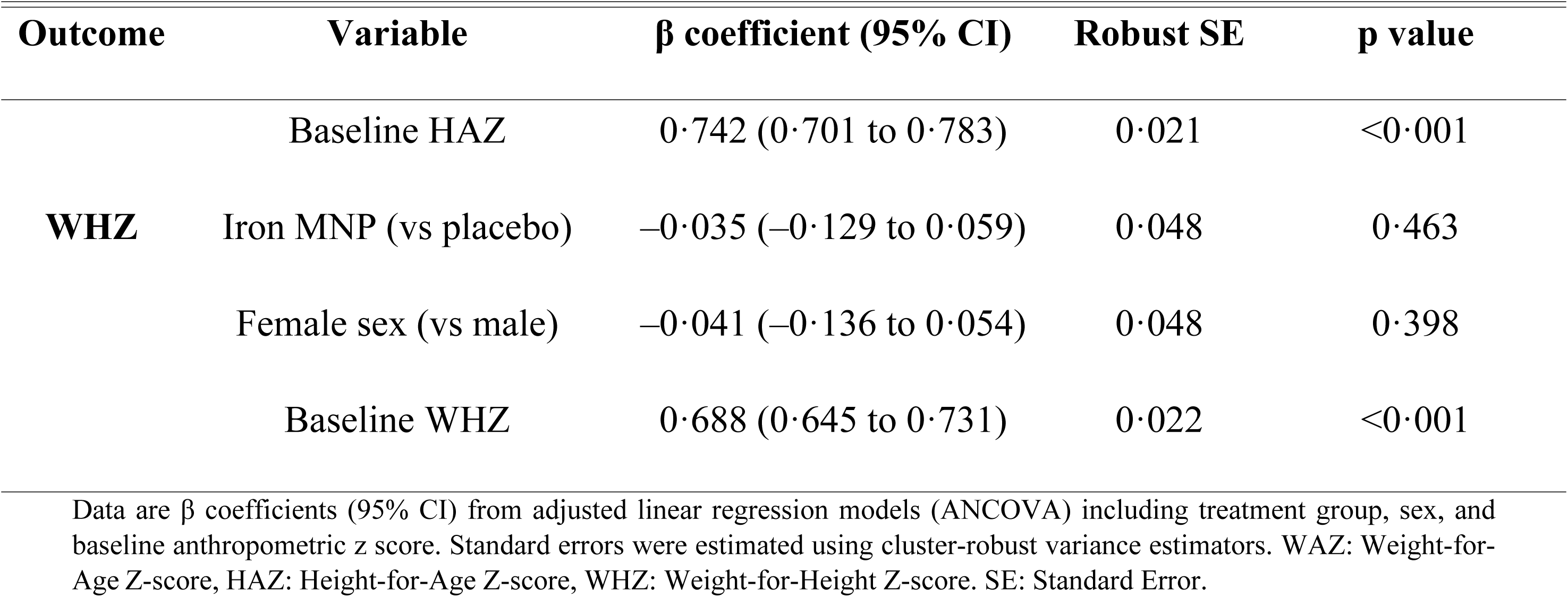
Adjusted association between intervention and anthropometric outcomes.

Mean baseline wasting prevalence was slightly higher among boys than girls in both groups. In the iron MNP group, boys had a baseline wasting prevalence of 9.6% compared with 8.9% among girls; in the placebo group, the corresponding figures were 8.5% and 5.7%, respectively. Two-sample t-tests showed no significant difference in wasting between the iron MNP and placebo groups among boys (p=0.573). However, a marginal difference was observed among girls (p=0.064), suggesting a possible sex-related trend, with girls in the iron MNP group having slightly higher baseline wasting than those in the placebo group.

The association between sex and anthropometric outcomes was also examined in the adjusted ANCOVA models (Table 2). Female sex was non-significantly associated with WAZ at endline (β = 0.028; 95% CI: –0.054 to 0.110; p = 0.503), indicating that boys and girls had similar weight-for-age outcomes after accounting for baseline nutritional status and treatment group (Table 2). Conversely, female sex was significantly associated with higher HAZ values than those of males (Table 2). The estimated coefficient was β = 0.149 (95% CI 0.045 to 0.253; p = 0.005), indicating that, on average, girls had higher linear growth scores than boys at endline, after adjusting for baseline HAZ and treatment assignment (Table 2). This finding implies that boys in the study population may have been more vulnerable to growth faltering than girls during the study period. Regarding weight-for-height z score, female sex was again non-significantly associated with WHZ (β = –0.041; 95% CI –0.136 to 0.054; p = 0.398), indicating no meaningful difference in acute nutritional status between boys and girls (Table 2).

### Distribution of Regression Residuals Across Models and Groups

Regression diagnostics indicated that key model assumptions were met (Figure 3). Residual-versus-fitted plots showed a random, evenly dispersed pattern around zero, with no discernible trends or curvature, indicating appropriate model specification and supporting the assumption of linear relationships. There was no evidence of residual clustering by treatment group, suggesting that model performance was consistent across the iron MNP and placebo arms (Figure 3). The normal Q–Q plots indicated that residuals were approximately normally distributed, with only minor deviations at the tails, commonly observed in field-based data and unlikely to meaningfully affect statistical inference. Scale–location plots revealed a relatively uniform spread of residuals across fitted values, with no clear signs of heteroskedasticity or funneling. Collectively, these findings indicate no substantial violations of regression assumptions and reinforce the reliability of the estimated intervention effects across study groups (Figure 3). The estimated intracluster correlation coefficients were low to moderate across outcomes, indicating limited but non-negligible within-cluster similarity (HAZ ICC=0.07; WAZ ICC=0.06; WHZ ICC=0.05). Corresponding design effects ranged from approximately 1.18 to 1.25, suggesting a modest inflation of variance due to clustering. These values indicate that although clustering was present, it did not substantially compromise statistical efficiency, and the use of cluster-adjusted standard errors was appropriate (Figure 3).

**Figure 3:**
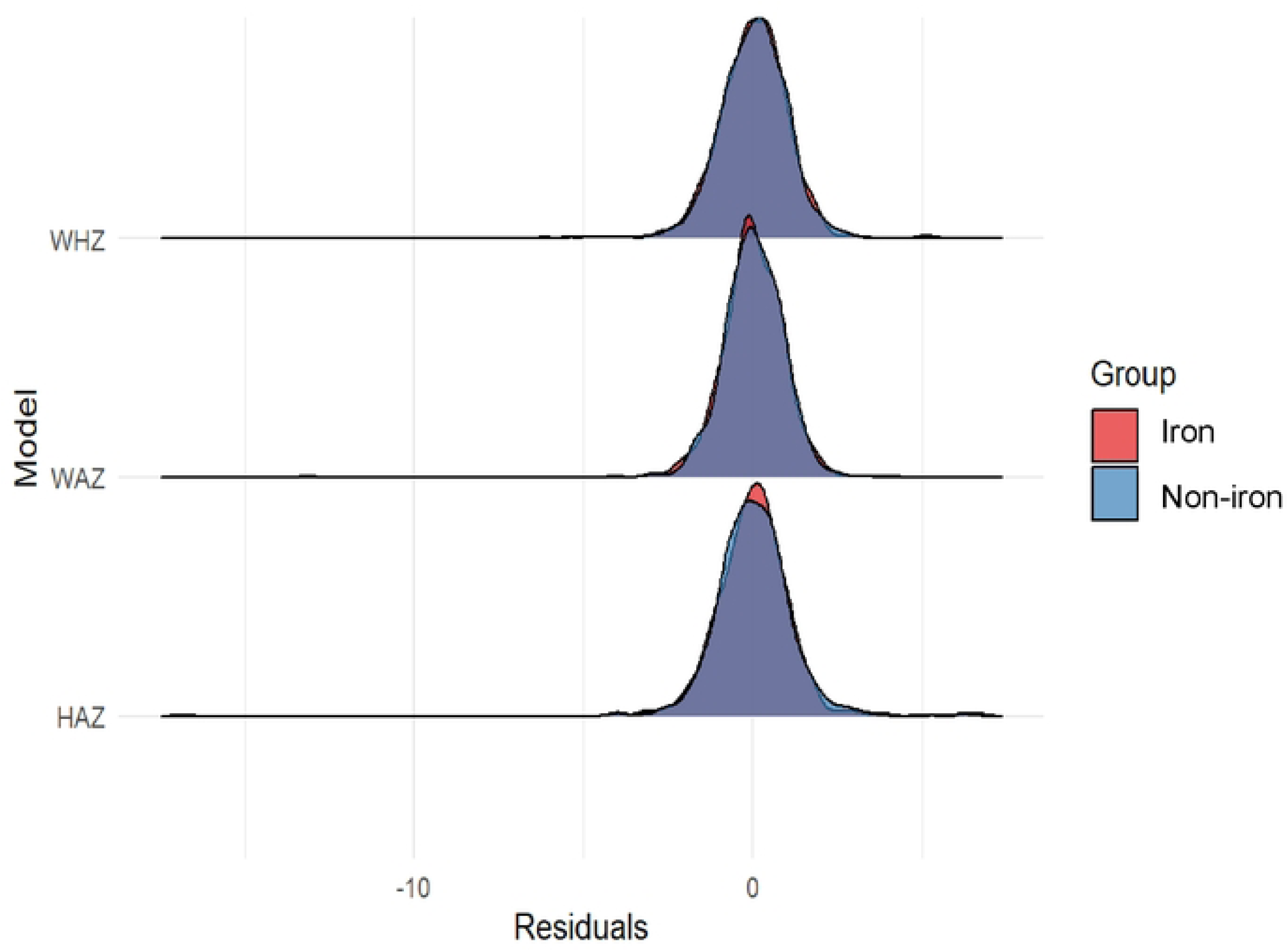
Distribution of Regression Residuals Across Models and Groups.

### Risk of Malaria-Induced Anaemia

The prevalence of malaria-induced anaemia at endline was lower in the iron MNP group. A binomial generalized linear model showed that the iron MNP group had a 5.0 percentage point lower risk of malaria-induced anaemia than the placebo group (Risk Difference = −0.050; 95% CI: −0.096 to −0.004; p=0.032) (Figure 4). Despite this reduction, a significant burden of anaemia remained in both groups. However, sex was not a significant predictor in this model (Figure 4).

**Figure 4:**
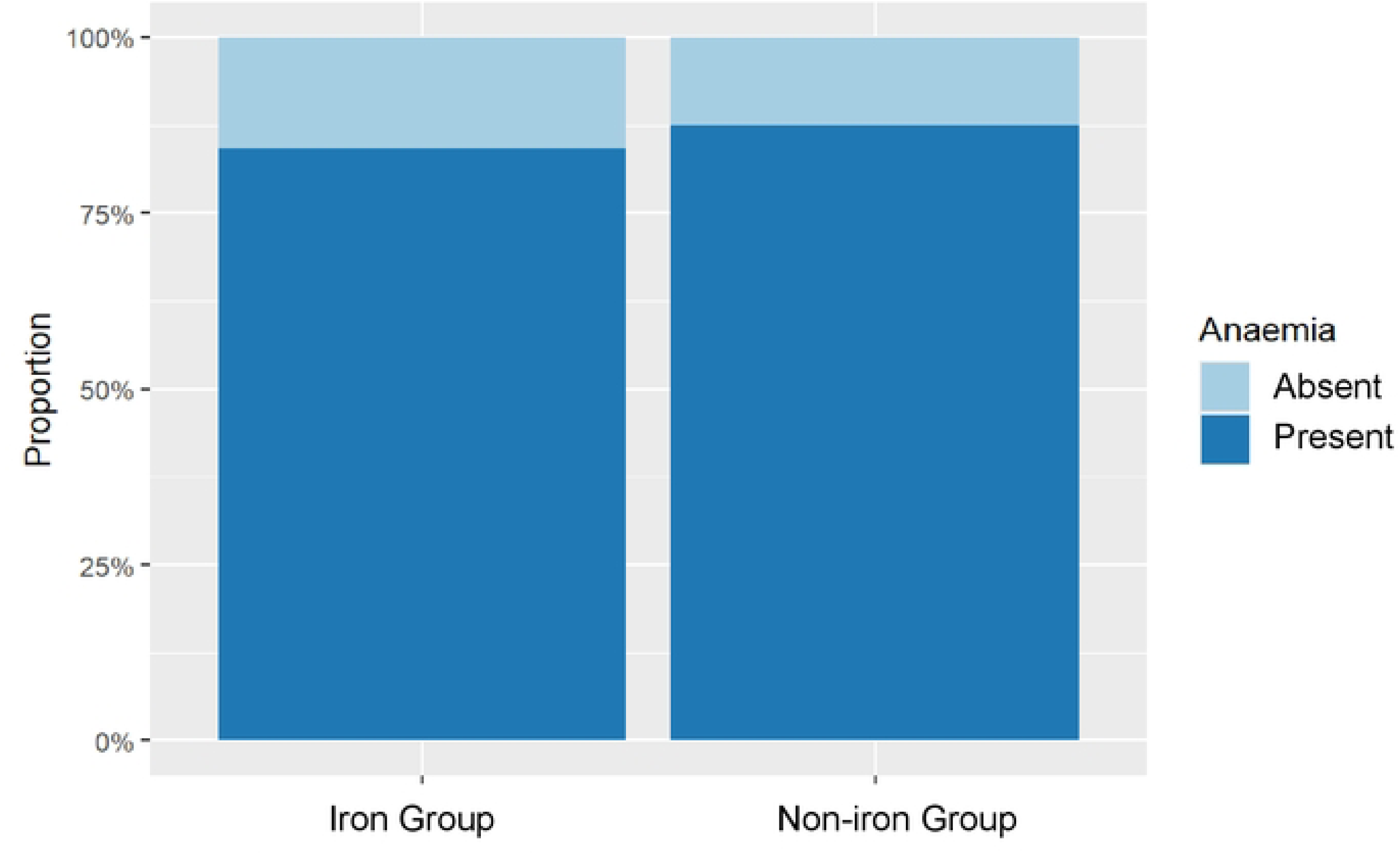
Anaemia Prevalence by Group.

## Discussion

This retrospective study analyzed secondary data from a large randomized cluster trial conducted in a high-malaria-transmission setting in Ghana. It found that daily prophylactic iron fortification with MNPs over five months did not improve linear or ponderal growth in preschool children, but was associated with a significant reduction in malaria-induced anaemia, as previously reported elsewhere (15–17).

The lack of impact on growth indicators aligns with previous findings in similar settings (16, 17). The troubling rise in stunting prevalence in both groups highlights that short-term micronutrient interventions alone are insufficient to reverse chronic malnutrition, which stems from complex, persistent factors such as recurrent infections, inadequate diets, and environmental stressors. However, in the adjusted sex-difference models, sex was significantly associated with higher HAZ scores in females than in males (Table 2). Thus, the estimated coefficient (β = 0.149 (95% CI 0.045 to 0.253; p =0.005)) indicates that after adjusting for baseline HAZ and treatment assignment, girls exhibited, on average, higher linear growth scores than boys at endline (Table 2). Nabwera and colleagues reported that integrating sensitivity and specific nutritional interventions halved malnutrition rates, though significant growth faltering remains (21). Tam and his team also reported that paediatric stunting and underweight improved with lipid-based nutrient supplementation (LNS), though MMN supplementation also marginally increased length-for-age z-scores (22). On the contrary, Mohammed and colleagues observed that age and breastfeeding duration were independently associated with linear growth and Hb, suggesting that linear growth and Hb are multifactorial, and implicating both nutritional and non-nutritional factors (23). Nonetheless, the notable sex effect on linear growth warrants further exploration of possible biological or caregiving differences.

The key finding of a protective effect against malaria-induced anaemia is both clinically and programmatically significant. These findings should be interpreted in the context of earlier evidence, particularly the Pemba trial, which reported increased adverse outcomes in settings with limited malaria control. (9, 22). Taken together, the current evidence indicates that the safety of iron interventions is context-dependent, varying according to formulation, dosage, baseline iron status, and the availability of effective malaria prevention, diagnosis, and treatment services (24). The use of microencapsulated ferrous fumarate in our study may have contributed to its safe profile by reducing the postprandial increase in non-transferrin-bound iron (NTBI), which can promote the growth of the malaria parasite (23). This supports the WHO stance that iron fortification (unlike high-dose supplementation) can be safe in malaria-endemic regions when implemented appropriately (22). The duration of the intervention (five months) may have been insufficient to detect meaningful changes in linear growth, which typically require longer follow-up. Additionally, the absence of detailed dietary and environmental data limits the ability to account for other determinants of growth fully.

A key strength of this analysis is the rigorous assessment of regression model assumptions, which supports the robustness of the findings (Figure 3). Residual-versus-fitted plots showed no evidence of systematic patterns, indicating appropriate model specification and linearity of associations. Residual distributions were comparable across intervention and control groups, suggesting that model fit was consistent and that group-specific deviations did not bias estimated intervention effects (Figure 3). Normal Q–Q plots indicated approximate normality of residuals, with only minor deviations at the tails, which are unlikely to materially affect inference, particularly given the use of robust standard errors (25, 26). Similarly, scale–location plots showed no evidence of heteroskedasticity, indicating stable variance across predicted values and study groups. Together, these diagnostics indicate no major violations of regression assumptions and strengthen confidence in the validity of the reported associations.

The observed reduction in malaria-induced anaemia without an increase in malaria risk is biologically plausible. This observation was supported by Castberg and his colleagues, who validated this method and the importance of fibroblast growth factor 23 (FGF23) as an independent inflammatory iron biomarker, whose role is increasingly recognized but still under investigation (27, 28). Iron supplementation can increase circulating iron availability, particularly non-transferrin-bound iron (NTBI), which may promote parasite replication (29). However, the use of microencapsulated ferrous fumarate in this study likely limited rapid increases in bioavailable iron, thereby reducing the risk of enhancing parasite growth. In addition, inflammation-mediated regulation of iron through hepcidin may further restrict iron availability during infection, providing a protective effect (28, 30).

Our findings should be interpreted considering the study’s limitations. The analysis relied on secondary data, which restricted the inclusion of potential confounders such as dietary diversity and household food security. Although five months is sufficient to observe haematological changes, it may be too brief to detect significant anthropometric improvements (31). Conducted in 2010, the study’s relevance may be affected by subsequent advances in malaria control, such as increased ITN coverage and malaria vaccination, though the biological interactions examined remain pertinent. We did not account for the potential contribution of other infections, particularly soil-transmitted helminths and other parasitic or bacterial infections, which are common in similar settings and may independently influence anaemia, iron metabolism, and child growth. Helminth infections, for example, can contribute to chronic blood loss, inflammation, and impaired nutrient absorption, thereby modifying both baseline nutritional status and the response to iron-containing interventions. The absence of data on these co-infections may therefore have resulted in residual confounding and limits our ability to fully disentangle the observed effects of micronutrient supplementation from broader infectious disease burdens.

## Conclusion

This study demonstrates that in high-transmission settings, iron-fortified MNPs appears safe and may reduce malaria-induced anaemia and provide modest protection without increasing the risk of malaria. Regression diagnostics indicated no meaningful violations of key model assumptions, supporting the robustness of the findings across study groups. However, to address the ongoing issue of stunting, iron fortification should be incorporated into broader public health strategies that include nutritional interventions, gender, effective malaria control, improved water, sanitation, and hygiene (WASH), and broader social and economic development efforts.

## Contributors

ET, FK, OO, FU, SD, BA, KT, and EN contributed to the overall study design, development of the analysis framework, and preparation of the manuscript. ET led the harmonization and analysis of the secondary trial data under the supervision of KT and EN. ET and EN were responsible for the review, structuring, and management of the secondary database. ET and JK conducted the statistical analyses. All authors critically reviewed the manuscript and approved the final version for submission. The authors acknowledge the KHRC and the original trial investigators for generating the primary dataset.

## Data Availability

https://doi.org/10.6084/m9.figshare.19768321.v1

## Acknowledgements

The authors express their profound gratitude to the study infants, young children and their caretakers; the KHRC staff and the field team; the chiefs, elders, and opinion leaders from the study communities; the study health facilities; the entire Ghana Health Service (GHS) staff in Wenchi Health Municipal and Tain Health District Directorates; and the Institutional Ethics Committees of KHRC, GHS, the Hospital for Sick Children-Toronto, Canada, DSMB, Food and Drugs Authority (FDA), Ghana.

## Availability of Data and Materials

The datasets are available and support the conclusions of this manuscript. https://doi.org/10.6084/m9.figshare.19768321.v1

